# Multiplex Immunohistochemical Phenotyping of T Cells in Primary Prostate Cancer

**DOI:** 10.1101/2021.09.03.21262635

**Authors:** Busra Ozbek, Onur Ertunc, Andrew Erickson, Igor Damasceno Vidal, Carolina Gomes Alexandre, Gunes Guner, Jessica L. Hicks, Tracy Jones, Janis M. Taube, Karen S. Sfanos, Srinivasan Yegnasubramanian, Angelo M De Marzo

## Abstract

Most prostate cancers are “immune cold” and poorly responsive to immune checkpoint inhibitors. However, the mechanisms responsible for the lack of a robust anti-tumor adaptive immune response in the prostate are poorly understood, which hinders the development of novel immunotherapeutic approaches. In addition, most inflammatory infiltrates in the prostate are centered around benign glands and stroma, which can confound the molecular characterization of the anti-tumor immune response. We analytically validated a chromogenic-based multiplex IHC approach and performed whole slide digital image analysis to quantify T cell subsets from the tumor microenvironment (TME) of primary prostatic adenocarcinomas. We trained a classifier to quantify the densities of eight T cell phenotypes separately in the tumor epithelial and stromal subcompartments. As an initial application, we tested the hypothesis that PTEN loss leads to an altered anti-tumor immune response by comparing matched regions of tumors within the same individual with and without PTEN loss. Our main findings in carcinomas (benign removed) include the following: i) CD4+ T cells are present at higher density than CD8+ T cells; ii) All T cell subsets are present at higher densities in the stromal compartment compared to the epithelial tumor compartment; iii) most CD4+ and CD8+ T cells are PD1+; iv) cancer foci with PTEN loss harbored increased numbers of T cells compared to regions without PTEN loss, in both stromal and epithelial compartments; v) the increases in T cells in PTEN loss regions were associated with ERG gene fusion status. This modular approach can apply to any IHC-validated antibody combination, sets the groundwork for more detailed spatial analyses, can help preserve small tissue samples, and can complement single cell and spatial genomic approaches.

## Introduction

The immune response within the tumor microenvironment (TME) is critical for immune surveillance, immune evasion, response to treatment and drug resistance ^1,2^. In the prostate, there is growing evidence for a pro-tumorigenic immune milieu that facilitates cancer initiation and progression ^3–11^. Despite this, various immunotherapies, including checkpoint immunotherapy, have generally shown very limited efficacy in prostate cancer ^12,13^, underscoring our need to better understand the anti-prostate cancer immune response.

An important approach to characterize anti-tumor immune responses is to elucidate the cellular and spatial composition of immune cells therein. While the non-neoplastic benign regions of the human prostate commonly harbor inflammatory infiltrates that are highly heterogeneous in composition, extent and location^3,9,14^, most prostatic adenocarcinomas contain few “chronic” or acute inflammatory cell foci (CD8+ T-cell rich or otherwise) directly involving tumor cell areas ^3,9,14–16^ and are generally considered non-inflamed “cold” tumors, consistent with an immune desert model^17^.

This immune desert TME signifies a general lack of a robust adaptive anti-tumor immune response in prostate cancer ^9,15,18,19^. The mechanisms leading to this immune desert-like state are not clear, but recent work has provided a number of insights. For example, most prostate carcinomas harbor a low tumor mutational burden,^20,21^ consistent with a paucity of mutation-associated neoantigens (MANA) that can serve as strong adaptive immune cell targets^22^. Furthermore, most primary hormone naive prostatic adenocarcinoma cells express little or no PDL1, suggestive of a lack of an adaptive immune cell-induced upregulation of this key inhibitory ligand ^23,24^. That a low number of MANA may often account for the relative immune desert in prostatic adenocarcinomas is consistent with the finding that in a subset of mismatch repair deficient cases that harbor a very high mutational burden, there are markedly increased numbers of intratumoral CD8+ T cells ^25,26^.

It is also possible that in many prostate cancers an active adaptive immune response is being repressed. For example, CD8+ T cells obtained from prostatic tissues of prostate cancer patients commonly express PD1, suggesting that a lack of strong immune response may result in part because of T cell “exhaustion”^27^. Furthermore, FOXP3-positive T regulatory cells (Treg), which can dampen anti-tumor immune responses, often accompany CD8+ T cells in the prostate, even in cases in which neoadjuvant therapies elicited an increase in tumor-associated CD8+ cells^28^. In addition, other cell types, including myeloid derived suppressor cells and/or M2 macrophages may also actively suppress the adaptive immune response ^10,29^. Finally, prostate tumor cells may down-regulate, or fail to express, important MHC proteins required for antigen presentation which are needed for adaptive immune cell recognition ^30,31^.

In terms of molecular subtypes of prostate cancer, even in cases without mismatch repair defects, there is inter-case heterogeneity in T cell density that is associated with the type of somatic molecular alterations in the tumor cells. For example, recent studies using monoplex chromogenic IHC have shown that the extent of CD8+ T cell infiltrates in prostate cancer is higher in cases harboring *TMPRSS2-ERG* gene fusions ^32^, PTEN loss^32^, *TP53* mutations^33^, and *CDK12* mutations^34^. However, the immunological significance of these findings remain unclear since the increases in CD8+ cells were generally modest and were often associated with increases in FOXP3 positive regulatory T cells (Treg).

Despite these advances, there are still important knowledge gaps in terms of the extent and spatial patterns of infiltration of key immune cell types in clinically localized prostate cancer. For example, in order to better understand the lack of efficacy of current immune checkpoint inhibitors, we need to learn more about the co-expression of regulatory molecules, such as PD-1, in different T cell subsets spatially within the prostate. Studies to determine the fraction of T cells expressing various immune checkpoint targets require a multiplex approach^35^. While methods such as flow cytometry and mass cytometry are highly suited for multiparameter cell type characterization, they inherently lose spatial information between cell and tissue types present.

The issue of the spatial distribution between cell types in the TME is especially important in primary invasive adenocarcinoma lesions because of the potential presence of pre-existing inflammation in the benign prostate. While the epithelial cells within the tumors are generally not directly infiltrated by abundant immune cells^3,9,15,16^, as stated above, the non-neoplastic benign regions of the human prostate commonly harbor highly spatially heterogeneous collections of inflammatory infiltrates ^3,9,14^. During the process of invasion, the adenocarcinoma glands frequently infiltrate into benign regions and become spatially intermingled with benign (and often inflamed) prostatic glandular and stromal tissue ^9^.

This indicates that virtually all studies that use tissue disrupting methods on specimens obtained from fresh primary tumor lesions from prostatectomies or biopsies, followed by downstream immune cell characterization (including traditional and mass-based flow cytometry assays, tissue imaging-based mass cytometry, RNAseq and single cell RNAseq), have an inherent problem; one cannot know whether the isolated immune cells being studied were present in the region of harvested tumor tissue because of an active anti-tumor immune response, or, if these cells were already present prior to tumor formation in regions of inflamed benign tissues. In the latter case, they are likely to be unrelated to an anti-tumor response. This problem of admixed benign-centered inflammation confounding results is also a potentially important concern for IHC/*in situ* based immune cell characterizations if benign regions within tumor foci are not excluded.

Although emerging technologies now allow 30-100 proteins to be characterized simultaneously in tissue samples^36^, these methods are still not widely available and have extremely low sample throughput. In terms of more readily available intermediate-level multiplex methods (e.g. for 5-8 simultaneous targets), two main approaches are being employed^37^. The first is multiplex IHC based on covalent deposition of tyramide-conjugated fluorophores, coupled with fluorescent slide scanning and spectral unmixing ^36,38–40^. The second is iterative IHC staining using a single chromagen (AEC), with sequential rounds of antibody staining, whole slide scanning and antibody stripping/removal. Each whole slide scan is then subjected to image registration and fusion and subsequent image analysis ^41–43^. While each of these methods have advantages and disadvantages ^36^, one key advantage to the iterative AEC-based approach is that it allows ready visualization by pathologists for whole slide quality control (Q/C) for each antibody, as well as implementation using widely available brightfield whole slide scanners that can image slides quickly at high magnification^36^. Also, there is no need for fluorescent imaging, or the separation of spectrally overlapping fluorophores. In this manuscript we adapted the AEC-based approach to develop a combined multiplex chromogenic T cell and epithelial cell panel to quantify the density of T cell subsets in the same whole slide tissue sections (CD4+ and CD8+, as well as their PD1 status, and Treg). This allowed us to study the spatial proximity of these cells to the tumor cells and tumor-associated stromal compartments.

Somatic *PTEN* inactivation is common in high grade prostatic adenocarcinoma, is associated with disease progression^44^. Furthermore, PTEN loss has been linked to a lack of immune directed tumor cell killing or immune suppression in a number of tumor types ^45^, including prostate cancer ^22,46^, and a prior study using singleplex IHC showed an association between PTEN loss and altered densities of CD3, CD8 and FOXP3 positive T cells^32^. Therefore, as a first application of our multiplex strategy presented here, we asked whether PTEN loss in tumor cells is associated with an altered adaptive immune response. Recent findings indicate that underlying germline genetic variations are associated with an altered anti-tumor immune response ^47,48^. Here, we used a unique study design for matched pairwise analysis in which each patient served as their own control because we compared tumor regions with and without PTEN loss using cancer foci from the same patient and same location within the prostate; thus, differences in T cell densities between PTEN loss and PTEN intact regions from the same patient could not be driven by differences in environmental exposures or germline genetic variants.

## Materials and Methods

### Antibodies

The primary antibodies and conditions are listed in **Supplemental Table 1**.

### Immunohistochemistry

4 micron sections paraffin sections were baked on a hot plate at 60 °C degree for 10 minutes, dewaxed using Xylenes, rehydrated in a series of graded alcohols to distilled water, rinsed in distilled water with 0.1% Tween solution. Slides transferred to a plastic jar filled with a suitable retrieval solution for each antibody and subjected to microwaving at full power for 1 minute, followed by 15 minutes at power level 20. Antigen retrieval buffers (Citrate, Vector Labs, H-3300; or Target Retrieval Solution, DAKO/Agilent Santa Clara, CA,, S170084-2) were used as indicated in **Supplemental Table 1**. Slides were cooled at room temperature for 5 minutes, followed by 2 washes in TBST. Slides were then subjected to endogenous peroxidase blocking for 5 minutes (Dual Endogenous Enzyme Block, DAKO, S2003). Then, the primary antibody was applied and slides were incubated for 45 minutes at room temperature or overnight at 4 °C degree (incubation time varied depending on primary antibody; see **Supplemental Table 1**). All remaining steps for each round of staining were carried out at room temperature. Slides were rinsed with Tris buffered Saline with Tween (TBST) and secondary antibodies applied (Ultravision Quanto from Leica, Buffalo Grove, IL; or PowerVision+ from Leica; **Supplemental Table 1)** for 30 minutes at room temperature. Slides were washed with TBST. 3-amino-9-ethylcarbazole (AEC)(ImmPACT AEC, VECTOR Labs, Burlingame CA) was applied for 20 minutes followed by TBST wash. Counterstaining was performed with hematoxylin (Mayers, DAKO, Diluted 1:4). Slides were washed with tap water for 2 minutes and distilled water for 1 minute. Then slides were coverslipped with aqueous mounting media (VectaMount AQ, Vector H-5501). Immunohistochemistry for PTEN was performed on an adjacent or near section to that of the multiplex staining using a Ventana automated staining platform (Ventana Discovery Ultra; Ventana Medical Systems, Tucson, AZ) employing a rabbit anti-human PTEN antibody (Clone D4.3 XP; Cell Signaling Technologies, Danvers, MA).This assay is highly sensitive and specific for the presence of underlying PTEN somatic genetic biallelic inactivation ^49,50^.

### AEC Removing and Antibody Stripping

Slides were scanned with a 40x objective using Ventana DP200 (Roche Diagnostics) digital whole slide scanner. Each whole slide scanned image was given a unique ID, including antibody and round of staining. After scanning, slides were decoverslipped in TBST and stained slides were dehydrated in an alcohol gradient to 95% ethanol. Slides were incubated in 95% ethanol until no visible AEC reaction product remained. Then, slides were re-hydrated through a change in 70% ethanol and then to dH_2_O for 2 minutes. Slides were then placed in the appropriate retrieval solution, and irradiated in a microwave oven at full power for 1 minute, followed by 15 minutes at power level 20. Slides were cooled at room temperature for 5 minutes, followed by 2 times wash in TBST. The slides were subjected to the next round of antibody staining, beginning with the blocking step. Primary antibodies,secondary antibodies, and chromogenic detection were serially added in the indicated order and condition shown in **Supplemental Table 1**. We switched the species between staining cycles, which provided the opportunity to microwave twice before using the secondary antibody targeting the same species. To validate the efficacy of the stripping method, for each antibody we performed a secondary-only protocol and found that each of these antibodies, except anti-keratin 8, were readily removed (see **Supplemental Fig. 1** for example). Keratin 8 was used as the last antibody in the sequence. We found low levels of variability in staining extent and intensities after performing duplicate parallel staining on serial sections for each antibody.

### Image Analysis

Digital image analysis was carried out using HALO 3.1(Indica Labs) software. After uploading images, elastic registration was run for the batch of scanned images for each case. Elastic registration transform files were exported to use in the image fusion step. Color deconvolution files were created as “xml” files which included the RGB optical density values of both hematoxylin and chromogenic positive stain. Next we created a batch file indicating which images to deconvolve, destination file names, the registration transform to use, and the correct color deconvolution files. We then ran the batch file to create two pseudofluorescent image layers in “tiff” format for each stain, one for hematoxylin and one for positive stained cells. After this step, we imported the pseudo fluorescent images back to HALO, ran the second registration, and fused the images. We set the image resolution value for the fused image. The High-Plex FL algorithm was adapted to analyze the positive cells and the colocalization of the markers in the fused images. Thresholds of each stain were set up by a pathologist with the real-time tune window. User-defined cell phenotypes were created to make the algorithm quantify double, triple, or quadruple positive cells. Cell segmentation was performed with the help of multiple parameters including minimum nuclear intensity, nuclear contrast threshold, and nuclear and membrane segmentation aggressiveness. We used a multi-step approach to identify the nuclear and cytoplasmic areas. Besides hematoxylin, the other nuclear stains (p63, FOXP3) in the staining panel also helped to optimize the nuclear segmentation. Membrane segmentation was set up using multiple staining markers including CK8, PD-1, CD3, CD4, and CD8.

To assess the validation of the cell phenotyping algorithm we selected 8 random areas, including areas from each of the PTEN loss or intact regions, and different inflammatory cell densities inside the tumor and counted positive cell numbers by manual visual counting.

The HALO random forest classifier was used to train a classifier to segment epithelial versus stromal regions in the fused images while all multiplex channels were turned on. Specifically, the classifier was trained on the basis of the manual annotation of epithelial and stromal regions. Cytokeratin-8 staining was employed to help train the software at this step. A pathologist visually checked the classifier and iteratively improved any inaccurate classification by adding additional training examples. The classifier that was created for each case was added to the High-Plex FL algorithm.

Analysis of the tumor regions from PTEN loss or intact areas were performed using an adjacent or near-adjacent cut of the slides stained separately for PTEN. The PTEN stained slide was scanned and the image was registered with the fused multiplex image of that case. Annotations were first made in the image of PTEN staining and used as a guide to annotate the tumor regions of interest in the fused image.

### Statistical Analysis

Data were tabulated and analyzed in Stata 15.1 for Mac OS,except for the linear regression analysis shown in **Figure 5 and Supplemental Figure 2**, which were analyzed using RStudio Ver 1.1456.

## Results

### Validation Studies

We developed a sequential multiplex IHC panel targeting T cells and epithelial markers. We first optimized each antibody (**Supplemental Table 1**) using AEC as the chromogen as a monoplex assay. Using human tonsil tissue as a control we found the expected patterns of staining for each marker. To define the tumor epithelial compartment separately from the tumor stromal compartment, we included an epithelial cell marker (keratin 8 or CK8) that stains all benign and neoplastic prostatic epithelial cells. To facilitate the ability to recognize benign glands near and inside of carcinoma lesions, and to recognize intraductal carcinoma (considered in most cases to be intraepithelial spread of preexisting high grade invasive carcinoma)^51^, we included staining for p63, which specifically stains nuclei of benign basal cells.

After each antibody staining round, the whole slides were digitally scanned and image analysis was used to generate fused multichannel pseudo-colored images (8 pseudo-color channels from 7 antibodies plus hematoxylin) (**Figs. 1-3**). **Fig. 1** shows an example of an image including both the original chromogenic slide scan and the results obtained after pseudocoloring and image fusion for CD8 and hematoxylin (**Fig. 1b**) or for 4 pseudocolors (**Fig. 1c**; p63, CD8, CK8 and hematoxylin). **Fig 2** shows both the original chromogenic image along with its corresponding 2-pseudo-color deconvolution image for 6 of the 7 antibodies in the panel (all except anti-p63 since p63 is not present in this image since it was taken from a region of invasive carcinoma).

**Figure 1.**
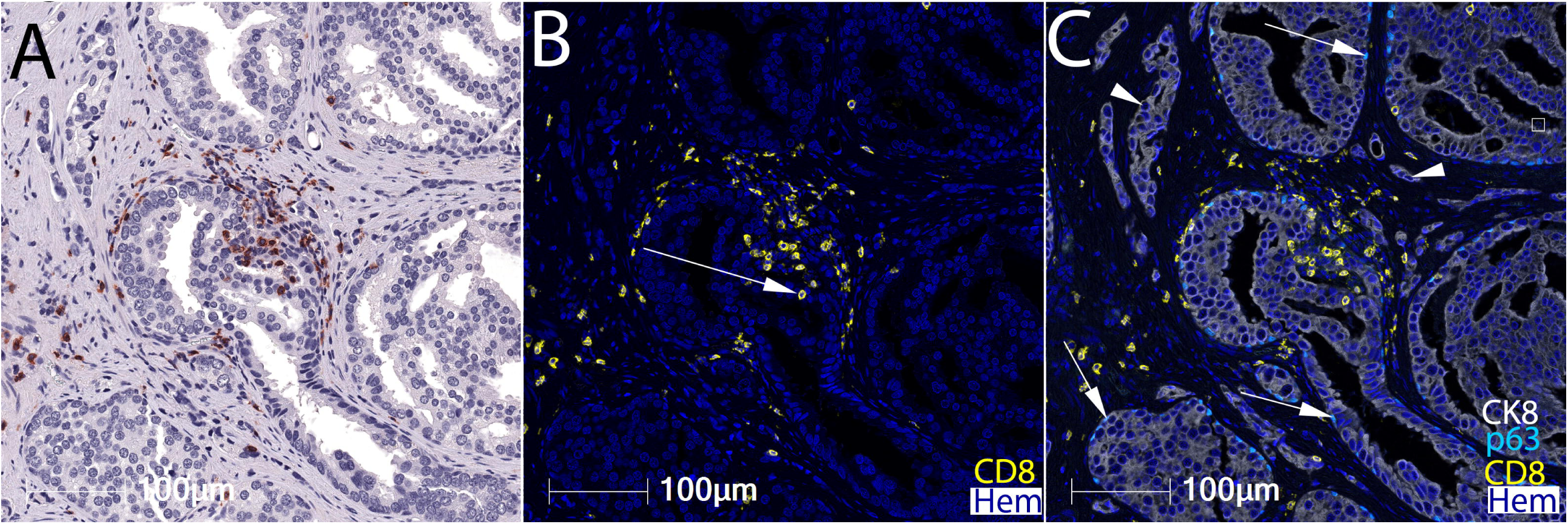
Chromogenic IHC for CD8 and cell segmentation using HALO. (**a**) IHC with anti-CD8 antibody on a prostate tissue section with AEC chromogen and hematoxylin counterstaining showing stromal and epithelial T cell infiltrates. (**a**) Shows only the CD8 stain, although this slide was stained with all 7 antibodies in the panel. (**b**) Pseudocolored image of region in (**a**) after color deconvolution in HALO showing CD8+ T cells in yellow and nuclei in blue. Hem indicates nuclei staining for hematoxylin. Arrow indicates intraepithelial CD8+ cell. (**c**) This shows additional channels from the multiplex pseudocolored image with keratin 8 shown in white, highlighting all epithelial cells, and with the p63 channel highlighting all basal cell nuclei in teal. The p63 reveals that some of the neoplastic glands have basal cells (arrows), which defines those glands as intraductal carcinoma. Arrowheads indicate invasive carcinoma in the stroma (non-intraductal). Note that the basal cells are not clearly evident in panel (**a**).

**Figure 2.**
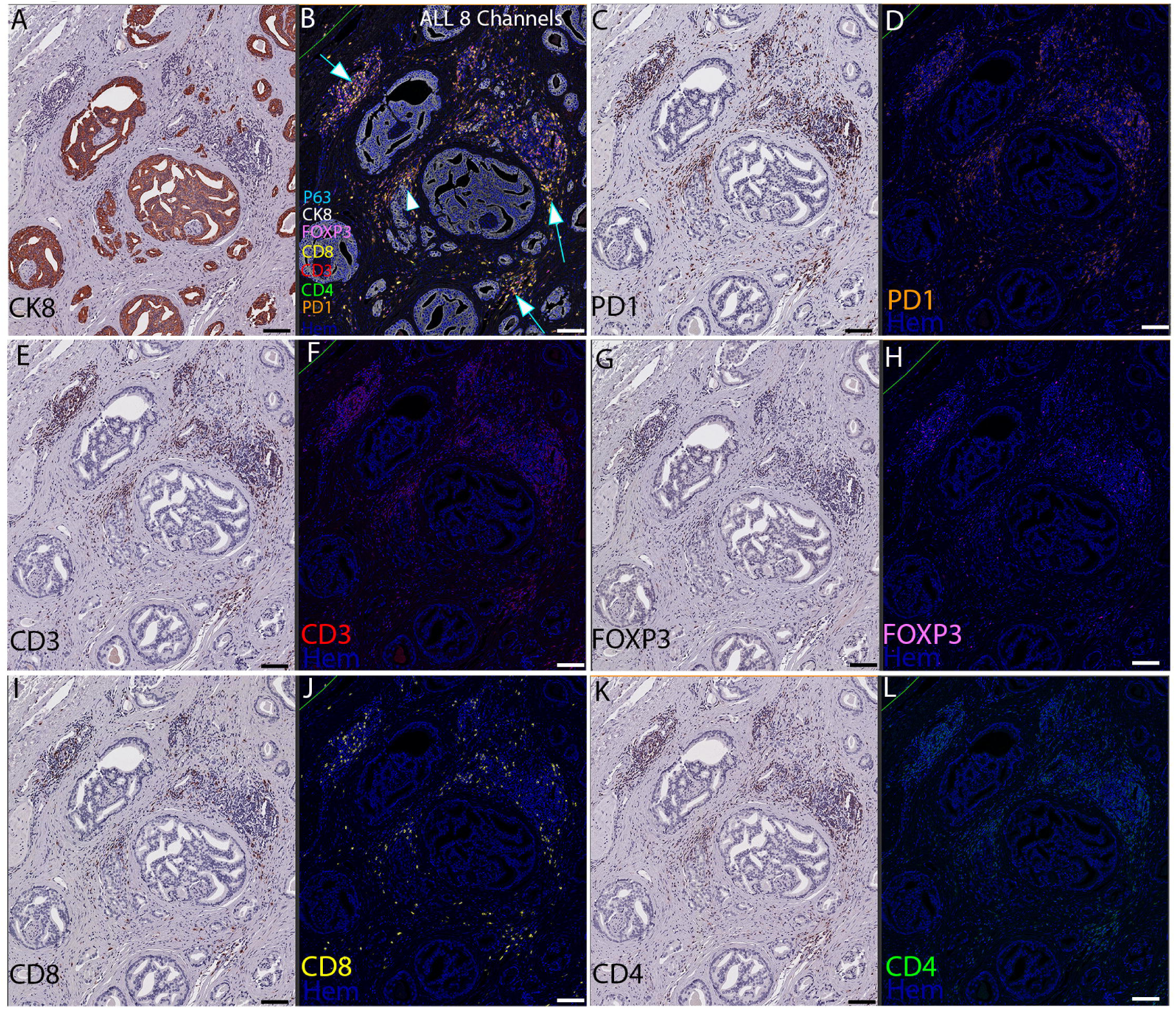
Chromogenic IHC with complete combined T Cell and epithelial panel. The region shown is of invasive carcinoma with no benign glands present. **a** Chromogenic IHC with anti-CK8 antibody. Note that all tumor glands are CK8 positive. **b** pseudocolored image showing all 8 channels (CD3/CD4/CD8/FOXP3/PD1/P63/CK8/Hematoxlyin) after image registration and fusion. Note that almost all inflammatory cells are present in tumor stromal compartment (arrows), with a few closely associated with tumor cells (arrowhead). **c** IHC with anti-PD1 antibody. **d** pseudocolored image of PD1 and hematoxylin channels only. **e** IHC with anti-CD3 antibody. **f** Pseudocolored image of CD3 and hematoxylin channels only. **g** IHC with anti-FOXP3 antibody. **h** Pseudocolored image of FOXP3 and hematoxylin channels only. **i** IHC with anti-CD8 antibody. **j** Pseudocolored image of CD8 and hematoxylin channels only. **k** IHC with anti-CD4 antibody. **l** pseudocolored image of CD4 and hematoxylin channels only. Original magnification x 100 for all panels (scale bar is 100 μm for each panel).

**Fig. 3** is an example of the 7-plex antibody plus hematoxylin panel (8 pseudocolors) in which 4 of the channels are shown (CD3, CD8, PD1 and hematoxylin) and where there is clear colocalization of multiple T cell markers on the same cell membranes for CD3, CD8 and PD1. This demonstrates that our image processing pipeline can precisely align successive whole slide scans at the individual cell/nucleus level. To calculate cell densities and spatial relationships of cell types of interest, the digitized image analysis data was used to segment cells into 8 distinct phenotypes (**Supplemental Table 1**) using the highplex fluorescent module in HALO.

**Figure 3.**
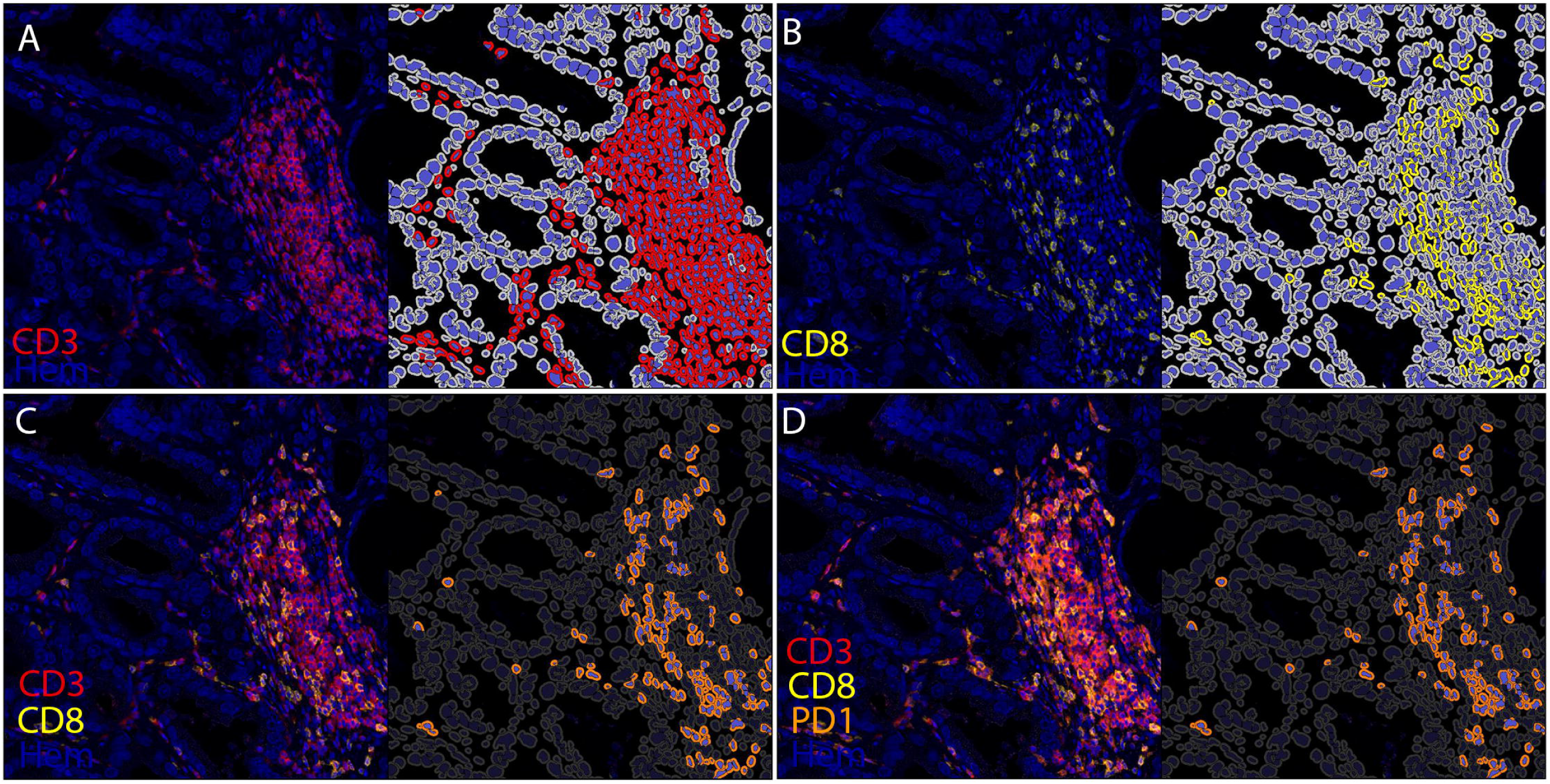
Cell segmentation for phenotyping. The figure shows 4 of the 8 pseudocolors for simplicity. Original magnification x 100. **a** The left panel shows a region of adenocarcinoma showing the pseudocolored images of CD3 (red) and hematoxylin (blue - Hem) and the right shows the segmentation of cells in HALO. On the right panel, CD3+ cells are segmented in which the nuclei are in blue and the cytoplasm/cell membranes are in red. Negative staining cells are shown with the nuclei in blue and cytoplasm/cell membranes in light gray. **b** The same region as that shown in (**a**) with CD8 (yellow) and hematoxylin (blue) pseudocolored image (left panel). On the right panel, CD8+ cells are segmented in which the nuclei are in blue and the cytoplasm/cell membranes are in yellow. Negative cells are shown as in (**a**) with the nuclei in blue and cytoplasm/cell membranes in light gray. **c** The left panel shows pseudocolored images of CD3 (red) and hematoxylin and CD8 (yellow) and the right shows segmentation for CD3+CD8+ double positive cells, with the nuclei in blue and cytoplasm/cell membranes outlined in orange. Negative cells have nuclei stained in blue and cytoplasm/cell membranes in dark gray. **d** Left panel shows pseudocolored images with CD3 (red), CD8 (yellow), PD1 (orange) and hematoxylin (blue) and right shows the phenotyped triple positive cells that are positive for CD3+CD8+PD1+ and they are colored as in (**c**) with the nuclei in blue and cytoplasm/cell membranes outlined in orange. Negative cells are colored as in (**c**) with nuclei stained in blue and cytoplasm/cell membranes in dark gray.

As previously reported^42^, we found that the sensitivity of staining was not substantially altered by the process of performing multiple rounds of staining and antibody stripping. To quantitatively address this issue in terms of enumerating specific cell phenotypes, we stained a tissue microarray (TMA) using the 7-plex antibody panel and separately stained serial sections with the same T cell binding antibodies by using monoplex chromogenic IHC performed by automated staining with DAB. Cell counts were obtained using the cytonuclear IHC module in HALO. There were strong correlations between the monoplex stained slides and the adjacently stained multiplex slides for CD3, CD8, CD4 and FOXP3 (**Fig. 4**), without a loss in cell number.

**Figure 4.**
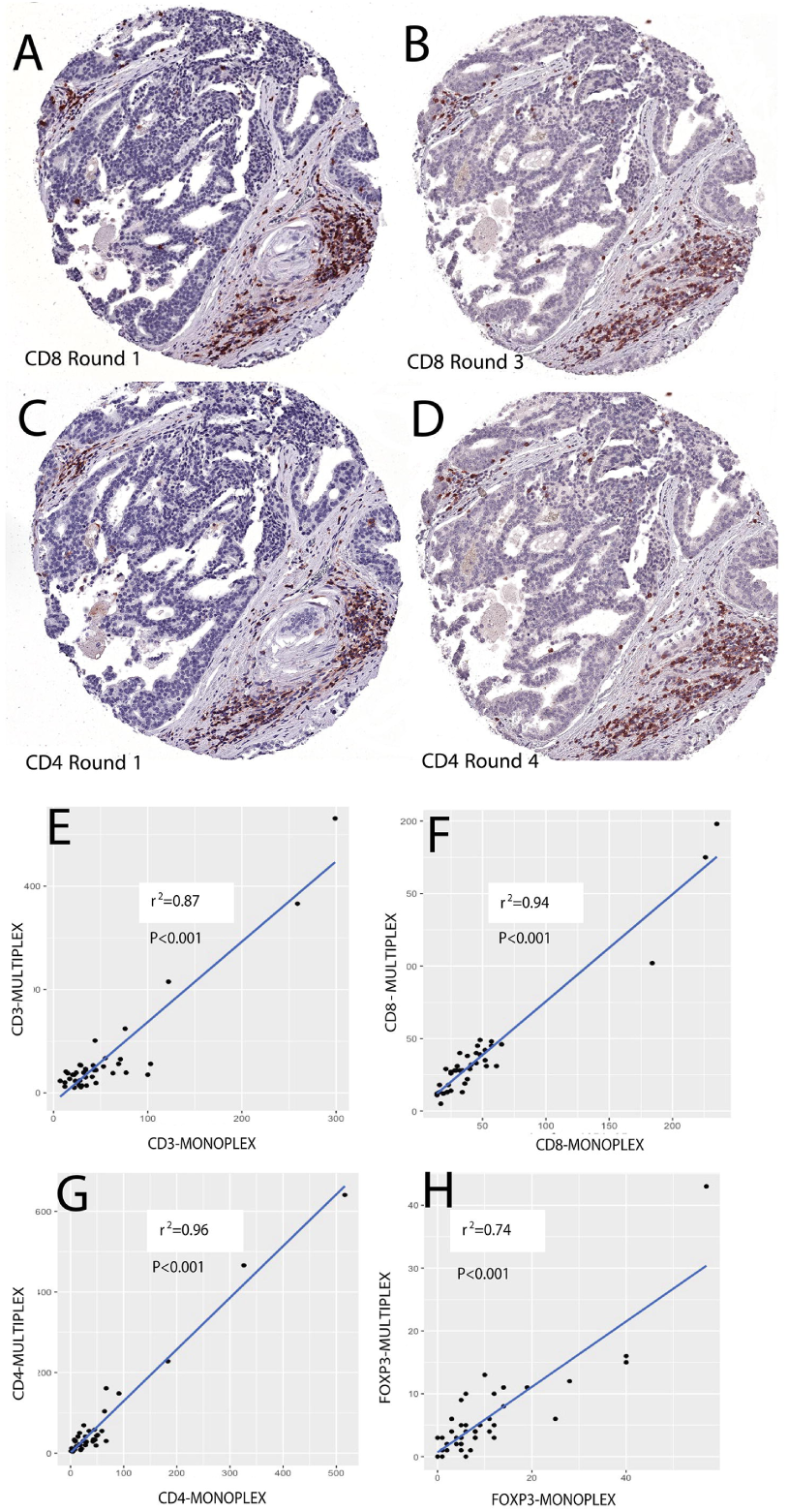
Validation of multiplex sequential assay by quantitative comparison to singleplex staining using a TMA. This shows an example of IHC staining of a TMA spot in the first round for CD8 (**a**) and in the 3rd round (**b**), and similarly for CD4 in the first round (**c**) and the 4th round (**d**). Note stromal T cells infiltrate on the lower right and upper left of images. **e-h** Stain-positive cells were counted manually within each TMA spot (each point on graphs represent cells counted in one TMA spot). Graphs show scatter plots separately correlating CD3+, CD4+, CD8+ and FOXP3+ cells using multiplex IHC (Y Axis) as compared with counts from adjacent slides stained using monoplex chromogenic IHC (X axes). TMA spots were analyzed using the cytonuclear IHC module in HALO. *P-*values and r^2^ are shown (simple linear regression).

To determine the accuracy of cell segmentation, phenotyping, and quantification by image analysis, we compared HALO cell quantification to manual cell counting for a number of regions of interest using standard slides from prostatectomies, and there was a strong correlation between these quantification methods (**Supplemental Fig. 2**).

Prior studies have suggested that, in addition to expression on T cells, CD4 is also present on a number of circulating monocytes^52^ and can be found in a subset of macrophages in porcine tissue^53^. We observed cells staining positively for CD4 that had a dendriform morphology consistent with that of some tissue macrophages. To examine this further, we performed a sequential multiplex panel using CD4, CD3, and CD163 (a macrophage marker) and identified a small but variable number of CD4+ cells that were also positive for CD163 and negative for CD3, indicating that there are CD4+ macrophages in human prostate tissue (not shown). Therefore, to avoid the possibility of counting CD4+ macrophages as T cells in our cell phenotyping, we used CD3 colocalization with CD4 to enumerate CD4+ T cells. We similarly used colocalization of CD3 and CD8 to enumerate CD8+ T cells, and CD3+CD4+FOXP3+ triple positive cells to enumerate Tregs.

### Quantification of T Cell Types in Prostate Cancer

#### Inflammatory Infiltrates are often Centered Around and in Benign Prostatic Glands

**Fig. 5** shows an example of the 7-plex/8 pseudocolor T cell panel showing a region of tumor that is infiltrating in and between benign glands. This region shows chronic inflammation in which the predominant spatial localization of the labeled T cells is centered in and around these benign glands/acini in a periglandular and intraepithelial location, with very few T cells closely associated with the adenocarcinoma glands. To accurately measure the phenotypes, number, and spatial relations of tumor infiltrating lymphocytes (TILs) in primary prostate cancers, our approach was to carefully annotate regions of tumor and to exclude regions of benign glands as much as possible inside and near the tumor (as seen by circled glands in **Fig. 5A**). **Supplemental Fig. 3** shows an additional set of examples at higher magnification in which benign glands were annotated for exclusion within the combined tumor epithelium and tumor stroma (“all tumor”). These highly annotated tissues are here-after referred to as Benign-Eliminated Tumor Regions of Interest (BET-ROIs). We also excluded regions with obvious staining artifacts (e.g. luminal contents staining positively in tumor regions).

**Figure 5.**
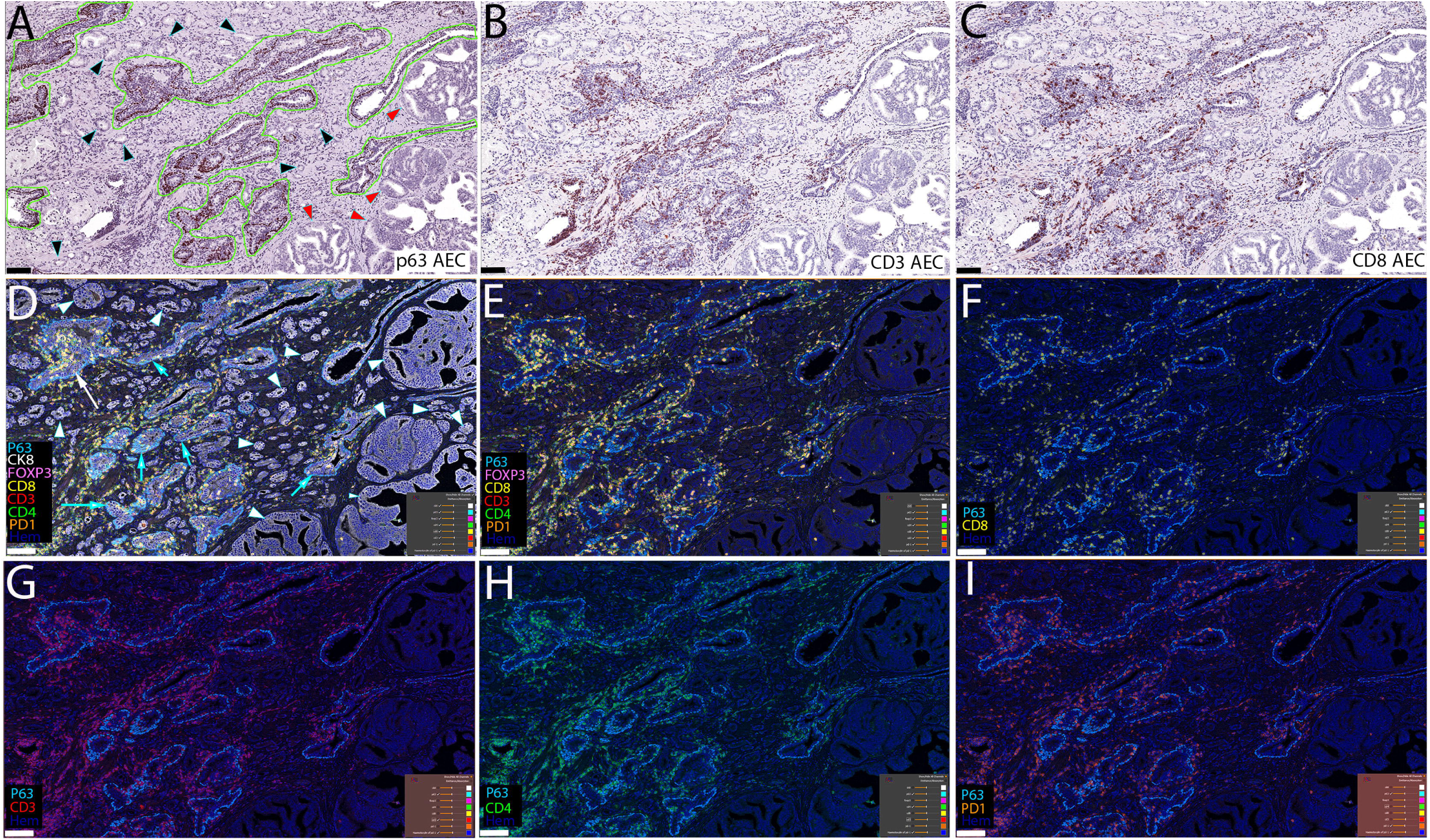
Lymphocytic infiltrates centered around benign glands in prostatic adenocarcinoma lesion (region of “tumor”). **a** Chromogenic IHC using AEC (red/brown chromagen) with anti-p63 antibody. Benign glands have a ring of p63 positive basal cell nuclei and are circled in green. Note small adenocarcinoma glands (black arrowheads) infiltrating between and around benign glands. Note larger cribriform adenocarcinoma glands on right (red arrowheads). The width of the entire field in this image is 2.1 mm and the height is 1.3 mm (original magnification x 100). **b** Chromogenic IHC of the same slide stained with anti-CD3 after stripping and removing prior chromogen and antibodies. Note the peri-glandular infiltrate of T cells around and within benign prostatic glands. **c** Chromogenic IHC with anti-CD8 of the same slide iteratively stained. **d** 8 pseudocolor image of the same slide after staining with all 7 antibodies and hematoxylin, shown after registration and fusion of all 8 channels in HALO. Arrows indicate benign glands and arrowheads indicate adenocarcinoma glands. Note benign glands outlined by p63 nuclear staining. **e** shows the same region as in (**d**), after removing only the keratin 8 channel. **f** This region is similar to (**d**) and (**e**) but only showing the CD8 layer along with p63 and hematoxylin. **g** This shows only the CD3, p63 and hematoxylin layers. **h** This shows only the CD4, p63 and hematoxylin layers. **i** This shows only the PD1, p63 and hematoxylin layers. Note the very low density of T cells near and directly involving carcinoma cells in this case, which is typical. **d-i** lower right thumbnail images transferred directly from HALO showing different channels selected. Scale bar is 100 μm for each panel.

To segment the tumor epithelial and stromal compartments separately for spatially resolved quantitative T cell density measurements, we used the CK8 staining to help create a tissue classifier in HALO, which determined separately the epithelial and stromal areas within the BET-ROIs (**Supplemental Fig. 4**). We then added this classifier to the Highplex-FL algorithm. The algorithm counted T cells as intraepithelial if more than half of the cell was inside the epithelium. If less than half of the cell was inside then it was counted as stromal.

### CD4 vs. CD8 in BET-ROIs and Epithelial Versus Stromal Compartments

For cell type specific quantification, we used a single standard slide representative of the index lesion from 15 radical prostatectomies (**Supplementary Table 2**). As a validation of our cell phenotyping and enumeration pipelines in HALO, within the BET-ROIs there was a strong linear correlation between the density of total T cells counted (all CD3+) and the sum of the densities of CD3+CD4+ double positive (referred to hereafter and in figures and tables as CD4+) plus CD3+CD8+ double positive cells (referred to hereafter and in figures and tables as CD8+) (r^2^= 0.96, *p* <0.001). In terms of T cell subsets, the tumors were infiltrated with a statistically significantly higher number of CD4+ cells than CD8+ T cells (**Fig. 6a**). The median density of CD8+ cells was 125 per mm^2^ (mean = 191) and of CD4+ was 315 cells per mm^2^ (mean = 317) (**Fig 6a**). The median percentage of total T cells was 38.9% for CD8+ and 61.1% for CD4+. Most of the CD8+ and CD4+ T cells were positive for PD1; there was a median of 83.5% (mean of 82.4%) of the CD8+ cells that were PD1 positive and a median of 75.6% (75.6%) of CD4+ cells were PD1 positive (**Fig. 6b**). The median density of CD3+CD4+FOXP3+ triple positive (Treg) cells was 32.8 cells per mm^2^ (mean = 39.1) and the median percentage of all CD3+ T cells that are Treg was 8.3%. The density of CD4+ cells correlated with the density of CD8+ cells and the density of each of these correlated with the density of Tregs (**Figure 7**). As expected from visual observations, there were significantly more cells in the tumor stromal compartment compared with the tumor epithelial compartment for each cell type (**Table 1)**. There was a significant correlation between the density in the stromal compartment and the density in the epithelial compartments for CD3+, CD8+ and Treg and a non-significant correlation for CD4+ cells. (**Figure 7D**).

**Table 1.** T Cell Densities in Epithelial and Stromal Tumor Compartments.

| Cell types |  | Epithelium | Stroma | *p* |
| --- | --- | --- | --- | --- |
| T cells all CD3+ | Median | 204.3 | 644.7 | 0.0007 |
|  | Mean | 214.9 | 787.9 |  |
|  | SD | 102.1 | 645.2 |  |
| CD8+ T Cells All | Median | 58.9 | 186.7 | 0.0007 |
|  | Mean | 71.3 | 308.4 |  |
|  | SD | 55.3 | 308 |  |
| CD4+T Helper cells All | Median | 98.4 | 323.5 | 0.0007 |
|  | Mean | 97.1 | 535.5 |  |
|  | SD | 42.6 | 401.7 |  |
| Regulatory T cells | Median | 9.6 | 49.1 | 0.0007 |
|  | Mean | 10.9 | 68.4 |  |
|  | SD | 6.8 | 64.2 |  |
| CD8+ T Cells - PD1 Positive | Median | 42.1 | 172.3 | 0.0007 |
|  | Mean | 54.3 | 260 |  |
|  | SD | 46.7 | 257.7 |  |
| CD8+ T Cells - PD1 Negative | Median | 18.6 | 37.2 | 0.0007 |
|  | Mean | 17 | 48.5 |  |
|  | SD | 11.2 | 52.9 |  |
| CD4+ T Cells - PD1 Positive | Median | 57.7 | 368.9 | 0.0007 |
|  | Mean | 55.3 | 427 |  |
|  | SD | 23.8 | 343.1 |  |
| CD4+ T Cells - PD1 Negative | Median | 32.9 | 99 | 0.0012 |
|  | Mean | 41.8 | 108.3 |  |
|  | SD | 29.8 | 66.7 |  |

*P*-values from Wilcoxon sign-rank test comparing densities in the tumor epithelium with densities in the tumor stroma.

**Figure 6.**
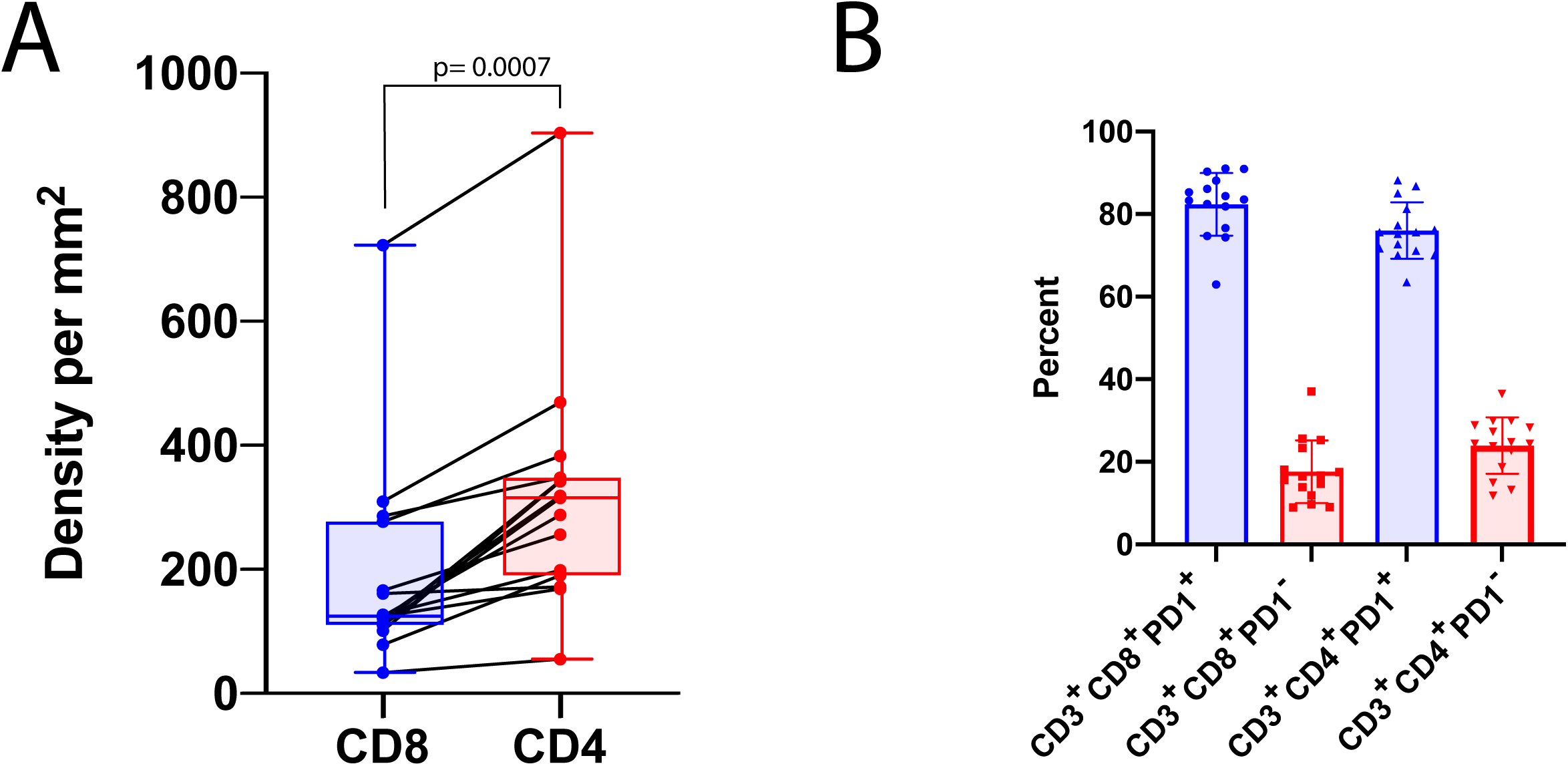
Higher density of CD4+ T cells than CD8+ T cells in BET-ROIs and the majority of both are PD1+. **a** The density of CD4+ T cells (CD3+CD4+) was significantly higher than CD8+ T cells (CD3+CD8+) (Wilcoxon signed-rank) in BET-ROIs when considering the combined epithelial and stromal tumor compartments together (all tumor). **b** The majority of both CD8+ T cells and CD4+ T cells in the all tumor area are PD1+. Each point on graphs represents one patient’s tumor region.

**Figure 7.**
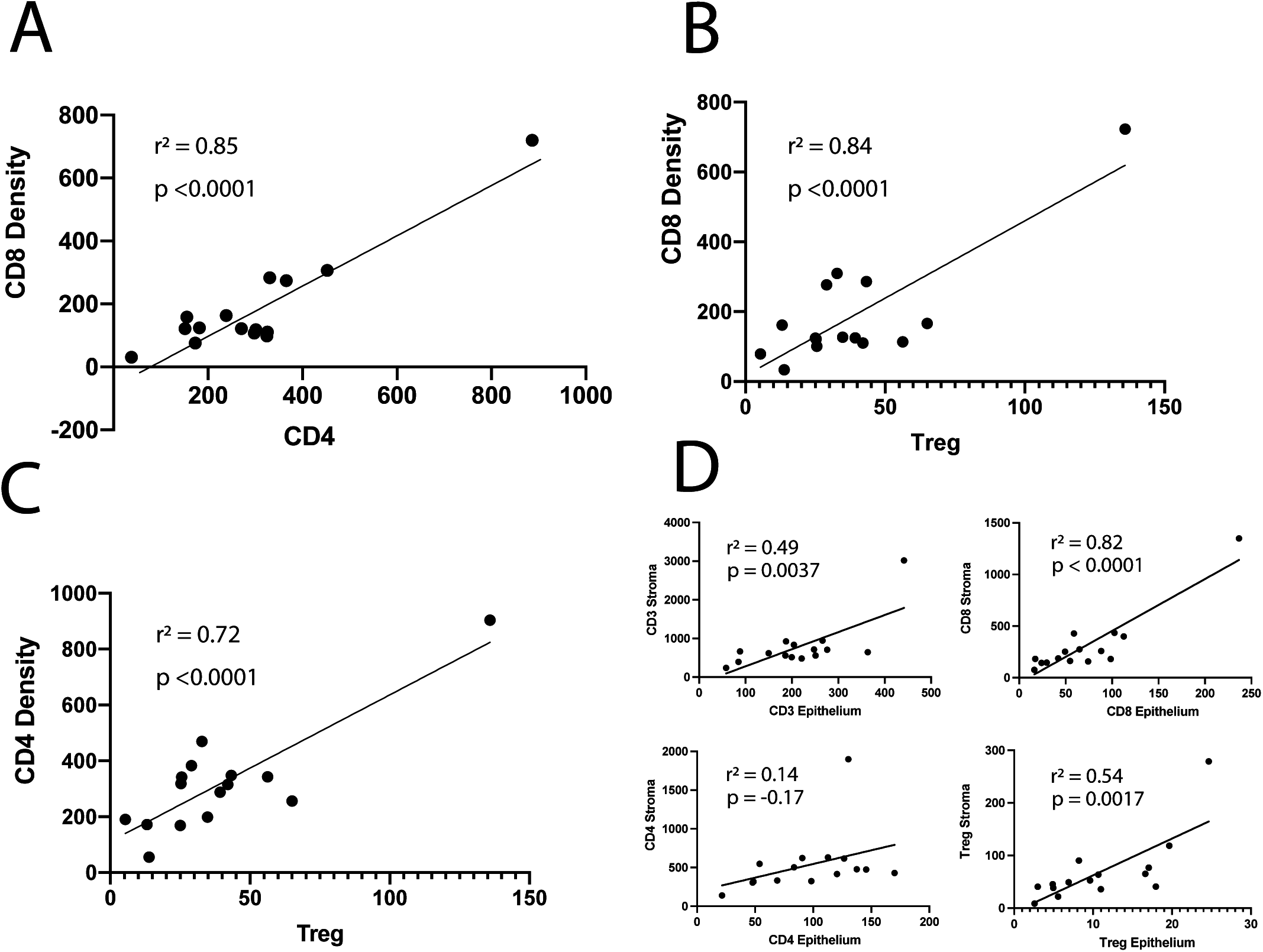
Correlation between CD4+, CD8+ and Treg epithelial densities overall, and epithelium compared with stroma in whole tumor regions. **a** CD4+ levels correlate with CD8+ levels. **b** CD8+ levels correlate with Treg levels. **c** CD4+ cell levels correlate with Treg levels. (**a-c**) All graphs show density per mm^2^ on the Y axis and results of simple linear regression analysis with r^2^ and p values shown. **d** The levels for total T cells (CD3+), CD4+ and CD8+ are all higher in stroma compared to epithelium, although the levels in stroma correlate with levels in epithelium.

### Relation to Molecular Subtype (PTEN and ERG Status)

PTEN protein loss as assessed by IHC staining is a robust measure reflecting PTEN genomic status of the tumor cells ^44,49,50^. In primary prostatic adenocarcinomas it is common to find individual tumor nodules that are heterogeneous for PTEN loss, indicating subclonal somatic genomic PTEN loss ^54–56^. To examine the association of T cell subtype densities with PTEN loss, each of the adenocarcinoma lesions used in this study was selected to harbor heterogeneous PTEN loss by IHC staining such that they contained separately annotatable regions that were intact for PTEN and regions with PTEN loss.This within-patient paired approach provides a unique opportunity to study the TME in relation to PTEN loss, whereby differences in environmental exposures or germline genetic variations that may affect gene expression, and hence the adaptive immune response to the tumor, have been controlled for. To increase the likelihood that the tumors with heterogeneous PTEN loss were related to each other clonally, all of them were located spatially adjacent to each other within a single FFPE block from the prostatectomies, and both components showed the same ERG status by IHC. Whole slide scans of the monoplex PTEN stained slides were used as a guide for the annotations of PTEN status on the multiplex pseudocolored images. **Fig. 8** shows an example of one of the cases showing heterogeneous PTEN staining. Using this approach for the combined epithelium + stroma tumor regions (all tumor) BET-ROIs, there were non-significant increases in total T cells (CD3+) and each major T cell subtype (CD4+, CD8+, Treg) in regions with PTEN loss, as compared with regions with intact PTEN **(Table 2**). These increases in regions of PTEN loss, while not readily apparent on visual microscopic inspection, occurred separately both in the tumor epithelial compartment and tumor stromal compartment, and whether the CD4+ or CD8+ T cells were PD1 positive or not (**Table 2**).

**Table 2.** T Cell Densities by PTEN Status for 8 Cell Phenotypes by Compartment (Regardless of ERG Status)

| Cell type | All Tumor = (Epithelium + Stroma) |  |  |  | Tumor Epithelium Only |  |  | Tumor Stroma Only |  |  |
| --- | --- | --- | --- | --- | --- | --- | --- | --- | --- | --- |
|  | PTEN Status |  |  |  | PTEN Status |  |  | PTEN Status |  |  |
|  |  | Intact | Loss | *p* | Intact | Loss | *p* | Intact | Loss | *p* |
| T cells all CD3+ | median | 430.1 | 438.8 | *0.0691* | 187.6 | 216.6 | *0.053* | 591 | 688.2 | *0.14* |
|  | mean | 387.9 | 562.2 |  | 177.8 | 252.9 |  | 599 | 854.2 |  |
|  | SD | 183.2 | 388.8 |  | 88.5 | 146.2 |  | 315.8 | 758.9 |  |
| CD8+ T Cells All | median | 127.8 | 136.7 | *0.173* | 49.3 | 57.6 | *0.91* | 190 | 215 | *0.112* |
|  | mean | 142.8 | 211.8 |  | 58.5 | 72.1 |  | 224.8 | 340.8 |  |
|  | SD | 91.1 | 205.7 |  | 39.3 | 65.3 |  | 141.5 | 373.3 |  |
| CD4+ T Helper cells All | median | 201.2 | 277.3 | *0.14* | 76.7 | 90.5 | *0.078* | 337 | 507 | *0.212* |
|  | mean | 246.4 | 346.9 |  | 80.6 | 107.6 |  | 411.8 | 571.2 |  |
|  | SD | 135.1 | 226.3 |  | 47.7 | 55.4 |  | 233.6 | 468.2 |  |
| Regulatory T cells | median | 25.8 | 38.8 | *0.0884* | 9.7 | 8.6 | *0.07* | 46.1 | 60.3 | *0.13* |
|  | mean | 32.4 | 44.6 |  | 8.9 | 13.9 |  | 57.1 | 74.6 |  |
|  | SD | 25.5 | 33.1 |  | 5 | 12.1 |  | 52.5 | 67.8 |  |
| CD8+ T Cells - PD1<sup>+</sup> | median | 107.4 | 116.8 | *0.125* | 35.3 | 48.3 | *0.69* | 168 | 175.1 | *0.09* |
|  | mean | 113.3 | 178.3 |  | 42.7 | 56 |  | 181.1 | 289.2 |  |
|  | SD | 72.9 | 178.4 |  | 29.7 | 56.4 |  | 111.4 | 318.3 |  |
| CD8+ T Cells - PD1<sup>-</sup> | median | 26.7 | 28.1 | *0.256* | 13.5 | 15 | *0.95* | 33.2 | 38.4 | *0.5* |
|  | mean | 29.5 | 33.5 |  | 15.7 | 16.1 |  | 43.7 | 51.6 |  |
|  | SD | 23.7 | 30.7 |  | 11.4 | 12.8 |  | 40.7 | 58.9 |  |
| CD4+ T Cells - PD1<sup>+</sup> | median | 177.7 | 215.4 | *0.1* | 38.2 | 57.1 | *0.23* | 277.1 | 407.1 | *0.14* |
|  | mean | 175.7 | 270.3 |  | 44.5 | 60.1 |  | 305.8 | 466.2 |  |
|  | SD | 88 | 203.4 |  | 22.6 | 40.3 |  | 156.7 | 417.9 |  |
| CD4+ T Cells - PD1<sup>-</sup> | median | 65.3 | 79 | *0.394* | 23 | 33.6 | *0.19* | 77.5 | 99.4 | *0.82* |
|  | mean | 70.7 | 76.7 |  | 36.1 | 47.5 |  | 106.1 | 105 |  |
|  | SD | 51.6 | 37.8 |  | 29.8 | 37.7 |  | 85.3 | 58.5 |  |

*P*-values reflect comparisons of PTEN intact vs. PTEN loss, Wilcoxon signed-rank test.

**Figure 8.**
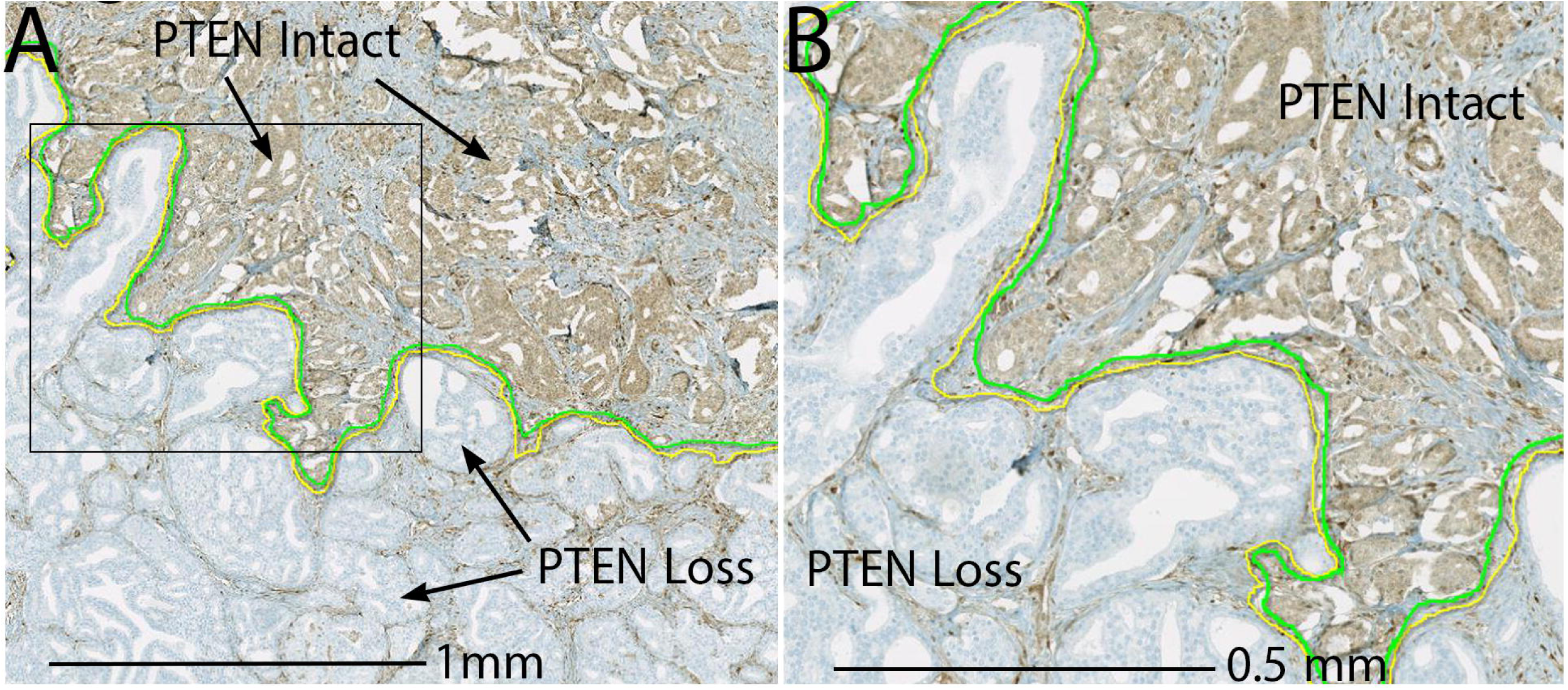
Adenocarcinoma with heterogeneous PTEN loss. Slide adjacent/near to the multiplex IHC stained slide showing IHC with anti-PTEN antibody staining (chromogenic with DAB). **a** Low power view with areas of intact PTEN staining and PTEN loss indicated. **b** Higher power view of boxed area in (**a**).

Since prostatic carcinomas with combined ERG and PTEN alterations have been associated with unique outcomes ^44^, we examined ERG expression in 13 of the 15 cases that had remaining available tissue; 7 cases were ERG positive (ERG+) and 6 were ERG negative (ERG-). As indicated above and as seen previously, the tumor nodules with heterogeneous PTEN loss were either homogeneously ERG positive or homogeneously ERG negative^54^. When stratifying by ERG status, in ERG+ positive cases there was a significant increase in Total T cells (CD3+), and Total CD8+ T cells in regions of PTEN loss compared to regions with intact PTEN (**Table 3**). There was also a significant increase in PD1+CD8+ cells in ERG positive cases in regions of PTEN loss, but not in PD1-CD8+ T cells (**Table 3**). Similarly, there was also an increase in PD1+ T CD4+ cells (CD4+PD1+) in PTEN loss regions in ERG+ cases but not in PD1-CD4+ cells (**Table 3**). There was a non-significant increase in PTEN loss regions of Treg in ERG+ and ERG-cases (**Table 3)**. Taken together, these findings indicate that increases in the density of specific T cell subtypes in regions of prostate carcinoma with PTEN loss, as compared with PTEN intact regions, are associated with ERG status of the underlying tumor. Furthermore, these increases in CD8 and CD4 positive cells in PTEN loss regions occur to a significant extent only in their respective PD1-positive cell fractions.

**Table 3.** Cell Density by PTEN Status for 8 Cell Phenotypes Stratified by ERG Status.

| Cell types |  | ERG Positive |  |  | ERG Negative |  |  |
| --- | --- | --- | --- | --- | --- | --- | --- |
|  |  | PTEN |  |  | PTEN |  |  |
|  |  | Intact | Loss | *p* (PTEN)* | Intact | Loss | *p* (PTEN)* |
| T Cells all CD3+ | median | 451.8 | 550 | 0.043 | 410.2 | 398.7 | 0.463 |
|  | mean | 403.8 | 719.4 |  | 343.1 | 425 |  |
|  | SD | 207.8 | 504.5 |  | 163 | 214 |  |
| CD8+ T Cells All | median | 114 | 162 | 0.043 | 126.7 | 129.6 | 0.92 |
|  | mean | 153 | 300 |  | 111 | 119.7 |  |
|  | SD | 113 | 276.1 |  | 53.1 | 61.2 |  |
| CD4+ T Cells All | median | 271.1 | 413.5 | 0.063 | 196.6 | 245.6 | 0.46 |
|  | mean | 286.6 | 493 |  | 187.9 | 219.5 |  |
|  | SD | 162.1 | 258 |  | 88.9 | 76.7 |  |
| Regulatory T cells | median | 25.6 | 43.3 | 0.09 | 23.7 | 39.4 | 0.7 |
|  | mean | 35.8 | 54.4 |  | 30.2 | 39.4 |  |
|  | SD | 28.6 | 42.7 |  | 28.4 | 24.1 |  |
| CD8+ T Cells - PD1 Positive | median | 87.8 | 121 | 0.03 | 112.2 | 113.3 | 0.75 |
|  | mean | 117 | 257 |  | 92.7 | 98.1 |  |
|  | SD | 85.7 | 237.7 |  | 42.6 | 45.8 |  |
| CD8+ T Cells - PD1 Negative | median | 27 | 30.4 | 0.5 | 14.4 | 16.3 | 0.46 |
|  | mean | 36.4 | 42.6 |  | 18.1 | 21.6 |  |
|  | SD | 30.3 | 40.7 |  | 13.7 | 18.1 |  |
| CD4+ T Cells - PD1 Positive | median | 199 | 290 | 0.043 | 162.4 | 186.5 | 0.6 |
|  | mean | 191.2 | 388.8 |  | 147.7 | 170.3 |  |
|  | SD | 105.4 | 244.1 |  | 67.6 | 83.2 |  |
| CD4+ T Cells - PD1 Negative | median | 72.1 | 95.4 | 0.5 | 34.6 | 41.6 | 0.35 |
|  | mean | 95.4 | 104.2 |  | 40.2 | 49.2 |  |
|  | SD | 59.6 | 29.6 |  | 26.3 | 24.5 |  |

\**P*-values reflect comparisons of PTEN intact vs. PTEN loss, Wilcoxon signed-rank test

When examining the data based on the ERG status of the case, without regard to PTEN, there were non-significantly higher densities of Total T cells (CD3+) and CD8+ T cells in the ERG+ cases (**Supplemental Table 3**). In terms of CD4+ T cells, there was a highly significantly elevated density in ERG+ compared with ERG-cases in CD4+ as well as both the PD1+ and PD1 - cell subtypes (**Supplemental Table 3**). There was a non-significant increase in Treg in ERG+ cases. Interestingly, when considering the PTEN status of the ERG+ tumors, the higher levels of CD4+ T cells (Total as well as PD1+ and PD1-), were statistically significant only in regions of PTEN loss (**Supplementary Table 4**). This finding supports a potentially important somatic genetic interaction between combined ERG+ and PTEN loss cases on CD4+ immune cell infiltrates. Treg density was increased non-significantly in ERG+ positive as compared with ERG-tumors, although unlike the other cell types, these increases appeared to be independent of PTEN status.

## Discussion

The detailed biological nature of the adaptive immune response in prostate cancer remains poorly understood^35^. This lack of knowledge hampers efforts for developing improved immunotherapeutic approaches for this disease. In the present study, we developed a chromogenic-based multiplex IHC approach to perform whole slide digital image analysis to quantify T cell subsets from primary prostatic carcinomas. We employed an image analysis pipeline to quantify 8 cell “phenotypes”, including total CD3+, total CD8+, total CD4+, Treg, CD8+/PD1+, CD8+PD1-, CD4+/PD1+, and CD4+/PD1-. We used a pan-prostate epithelial marker (anti-CK8) to train a classifier to quantify the densities of each of these T cell phenotypes separately in the epithelial and stromal subcompartments of the iTME.

We and others have recognized an important concern for studies that attempt to molecularly characterize and spatially map cells responsible for anti-tumor adaptive immune recognition and control (e.g. tumor infiltrating lymphocytes or TILs and other immune cells) in primary prostate cancers. That is, perhaps with the exception of rare high grade prostate cancers with mismatch repair deficiency ^25^, most immune infiltrates in prostates with or without cancer (either visibly evident by H&E staining or by IHC or in situ hybridization) are centered around and within benign glands and stroma ^6,9,15,16^. The implications of this long standing observation is that many of the T lymphocytes in prostates with cancer may be responding to benign-associated tissues/antigens, and may not be directed towards tumor-related antigens, which can confound the understanding of anti-tumor immune responses in the prostate. While this confounding is generally not taken into account, it should nevertheless be expected in studies that use cell dissociation after tumor harvesting (e.g. in flow cytometry and/or single cell RNAseq). In addition, this issue is also a potentially significant pitfall even in studies using *in situ* techniques (e.g. IHC, multiplex IHC [including highly multiplex approaches such as digital spatial profiling or imaging mass cytometry or CODEX for protein], and/or multiplex and highly multiplex *in situ* hybridization approaches for RNA) if investigators that do not attempt to very carefully annotate tissues to spatially exclude benign prostatic glands and stroma (**See Fig. 5 and Supp Fig. 2**). In this study, therefore, we carefully excluded benign areas, such that the quantified regions (BET-ROIs) were spatially enriched for carcinoma.

As a first proof of concept of our multiplex approach, we employed a unique study design by using paired adjacent PTEN-Loss/PTEN-intact tumors with both components harboring the same ERG status, to further investigate the hypothesis that PTEN loss is associated with an altered adaptive immune response. This approach provided an opportunity to control for underlying germline genetic variations that could affect the anti-tumor T cell response ^47,48^. Our main findings included the following: i) there are higher densities of all T cell subsets in the stromal compartment as compared with the epithelial tumor compartment; ii) there are higher numbers of CD4+ T cells than CD8+ T cells; iii) most CD4+ and CD8+ T cells are PD1+; iv) there are increased T cells and T cell subtypes in regions of PTEN loss in both stromal and epithelial compartments; v) the increases in specific T cell numbers in PTEN loss regions were related to the ERG gene fusion status of the cases.

A number of studies have examined the prognostic significance of CD8+ T cells in prostate cancer.^32,57,58^. However, the findings have generally been mixed and there is not a consensus regarding whether increased CD8+ T cells are related to an improved outcome or a worse outcome. Most prior studies on the prognostic significance of CD8+ in prostate cancer have not used a multiplex approach, and none have incorporated the PD1 status of the CD8 cells. Thus, it is not clear if the conflicting results relate to different study designs, the failure to carefully exclude benign areas in the quantification, or the lack of more in depth analysis of the cellular phenotypes with multiplex approaches. Our current approach in which we found that a high percentage of BET-ROI CD8+ T cells were PD1+, indicates that many, if not most, of the tumor-associated CD8+ T cells in prostate cancer may have an exhausted phenotype, as suggested previously ^16,27^. This, coupled with the correlation of CD8+ cells with FOXP3+ CD4+ Tregs (**Fig. 7B**), as also seen previously ^28,32^, supports the concept that perhaps more than one mechanism of adaptive immune suppression is active in the prostate cancer iTME ^35,59^.

The higher density of CD4+ cells, compared to CD8+ cells, in the BET-ROIs has been seen previously by Ebelt et al., although those studies did not employ multiplex staining, and did not use quantitative image analysis ^15,16^. In our study we used image analysis and also found that the higher levels of CD4+ cells, as compared with CD8+ cells, were present both in the stromal and epithelial tumor compartments of the TME. Furthermore, like the CD8+ T cells, most of the CD4+ T cells in the tumor lesions in the present study were PD1+. These results suggest that there is a need for additional studies of CD4+ T cell subsets in prostate cancer to better understand their functional significance. For example, Goods et al., recently found that PD1+ CD4+ cells lacking FOXP3 displayed lower proliferation compared with PD1-CD4+ cells and also showed a transcriptional signature suggestive of exhaustion^60^. The results also suggest that future work should include studies of the spatial relations between additional subsets of T cells with each other and with the tumor cells. By selection of other antibodies for multiplexing, our current approach, perhaps in conjunction with other approaches such as single cell RNAseq and *in situ* transcriptomics, is amenable to performing these analyses.

A number of prior studies have examined features of the adaptive immune response in prostate cancer in relation to specific somatic molecular alterations ^32–34^. In this study we focused on examining the relation to PTEN status. As seen previously, we found an increased number of CD8+ T cells in tumors with PTEN loss^32^, although ours is the first study to use PTEN negative and PTEN positive regions from the same patient’s tumors. We additionally report the novel findings that the increases were present in both the epithelial and stromal tumor compartments and there are also increases in total CD4+ T cells in tumors with PTEN loss. Interestingly, the effect of PTEN loss appeared to be modified by the ERG status of the tumors. For example, in ERG negative cases, there was either no increase with PTEN loss, or, any increases in any of the 8 phenotypes were non-significant. By contrast, in ERG+ cases, there were significant increases in total T cells (CD3+), and total CD8+T cells in PTEN loss regions compared with PTEN intact regions. Interestingly, for the CD8+ T cells, the increase in relation to PTEN loss was mostly found in the PD1+ subset, where the difference was significant. The increase in Treg in ERG+ cases in relation to PTEN loss was still not significant, nor was the increase in T (CD4+). As with CD8+ T cells, however, there was a significant increase in CD4+ T cells that were PD1+, but not PD1-.

When we consider the ERG status of the tumors by itself (**Supplemental Table 3**), the only significant increases in ERG+ compared with ERG-cases were in total T cells (CD4+), and this was significant in both PD1+ and PD1-subsets of these cells. Interestingly, the increase in CD4+ T cells in relation to the ERG status was present only in the PTEN loss regions. These findings are generally consistent with the increased CD3, CD8 and FOXP3 seen by Kaur et al. using singleplex assays and tissue microarrays, in which there were increases in all of these cell types with PTEN loss and the highest densities of each were found in cases with PTEN loss that were ERG+ primary untreated prostatic carcinomas with PTEN loss^32^. Taken together, these results suggest a complex interaction between PTEN and ERG in the prostate cancer iTME. In a number of studies, the adverse effects of PTEN loss on long term outcomes appears to be attenuated in ERG+ cases^44^ and this may relate to a modified adaptive immune response in these cases, although precisely how this could lead to an improved prognosis remains unclear. Additional studies with more refined analysis of the T cell subsets in such cases may provide further insights into these findings.

In terms of Tregs, a number of prior studies have evaluated the presence of FOXP3 positive cells in and around prostate cancer ^16,28,32,61^, and at least two prior studies found increased Tregs in prostate cancers with PTEN loss ^32,46^. Our results are generally consistent with the finding of increased FOXP3 positive cells in prostatic carcinoma with PTEN loss, although in the present study the increases were not statistically significant. Kaur et al., also found that the increase in CD8+ cells in prostatic carcinomas with PTEN loss was also accompanied by increased FOXP3 positive cells^32^, suggesting that any increase in T effector cells based on PTEN loss may be offset by similar increases in Treg. In the present study we found a similar relationship.

In terms of multiplex IHC studies in prostate cancer, while ours is the first to apply iterative chromogenic multiplexing, a number of other studies have employed fluorescence based techniques on primary prostate cancers ^58,62–64^. Like our study, all of these represent promising approaches, but also are limited by relatively low throughput. We do anticipate improved workflows for the current AEC-based approach in the near future. For example, during the preparation of this manuscript we upgraded our HALO software (from 3.0 to 3.2), and this results in a much faster image deconvolution and fusion processing pipeline. Coupled with future approaches using machine learning, we envision the ability to analyze multiplex-stained slides with greatly improved throughput in the future.

Our study has a number of limitations. First, given the relatively low throughput of the staining and analysis, we employed a total of only 15 RRP specimens. Additional studies with larger numbers of specimens are currently underway using tissue microarrays to better evaluate associations between the different T cell phenotypes studied here and long term outcome. Furthermore, our staining approach is not yet amenable to automated IHC staining, which will limit implementation currently. Finally, we only began to examine spatial relationships in terms of analysis of tumor epithelium and stroma and additional spatial studies (such as spatial relation between different T cell subtypes, and spatial relation between these and tumor cells) are planned.

In conclusion, we analytically validated an interactive chromogenic multiplex approach applied to quantify T cell subsets in primary prostate tumors using large tissue sections and whole slide image analysis. We report increased levels of a number of T cell subsets in the epithelial and stromal tumor compartments in prostatic carcinomas in regions of PTEN loss using a novel paired approach that controls for underlying germline genetic differences between patients. Since this multiplex approach is modular, this system is capable of being extended to employ additional panels to include any IHC-validated antibodies ^42^. Also, one can increase the number of markers in the same panel and this analysis pipeline is amenable to more in depth spatial analyses^65^. This approach is also amenable to prostate biopsies from either primary or metastatic tumors. Finally, the multiplex approach also helps to preserve precious tissue samples (e.g. small biopsies) and can complement emerging high content genomic single cell approaches using both tissue disruptions and *in situ* transcriptomics.

## Supporting information

All supplemental files with ages fixed in table

## Data Availability

All data will be made available.

## ACKNOWLEDGEMENTS

This study was supported by the NIH/NCI SPORE in Prostate Cancer (P50CA58236), and the NIH/NCI U01 CA196390 for the Molecular and Cellular Characterization of Screen Detected Lesions (MCL), the U.S. Department of Defense Prostate Cancer Research Program (PCRP) Prostate Cancer Biospecimen Network Site (W81XWH-18-2-0015), and The Johns Hopkins Sidney Kimmel Comprehensive Cancer Center Oncology Tissue Services Laboratory supported by NIH/NCI grant P30 CA006973.

