## Supplementary material for "Multiplex Immunohistochemical Phenotyping of T Cells in Primary Prostate Cancer": All supplemental files with ages fixed in table

**Supplemental Figure 1**

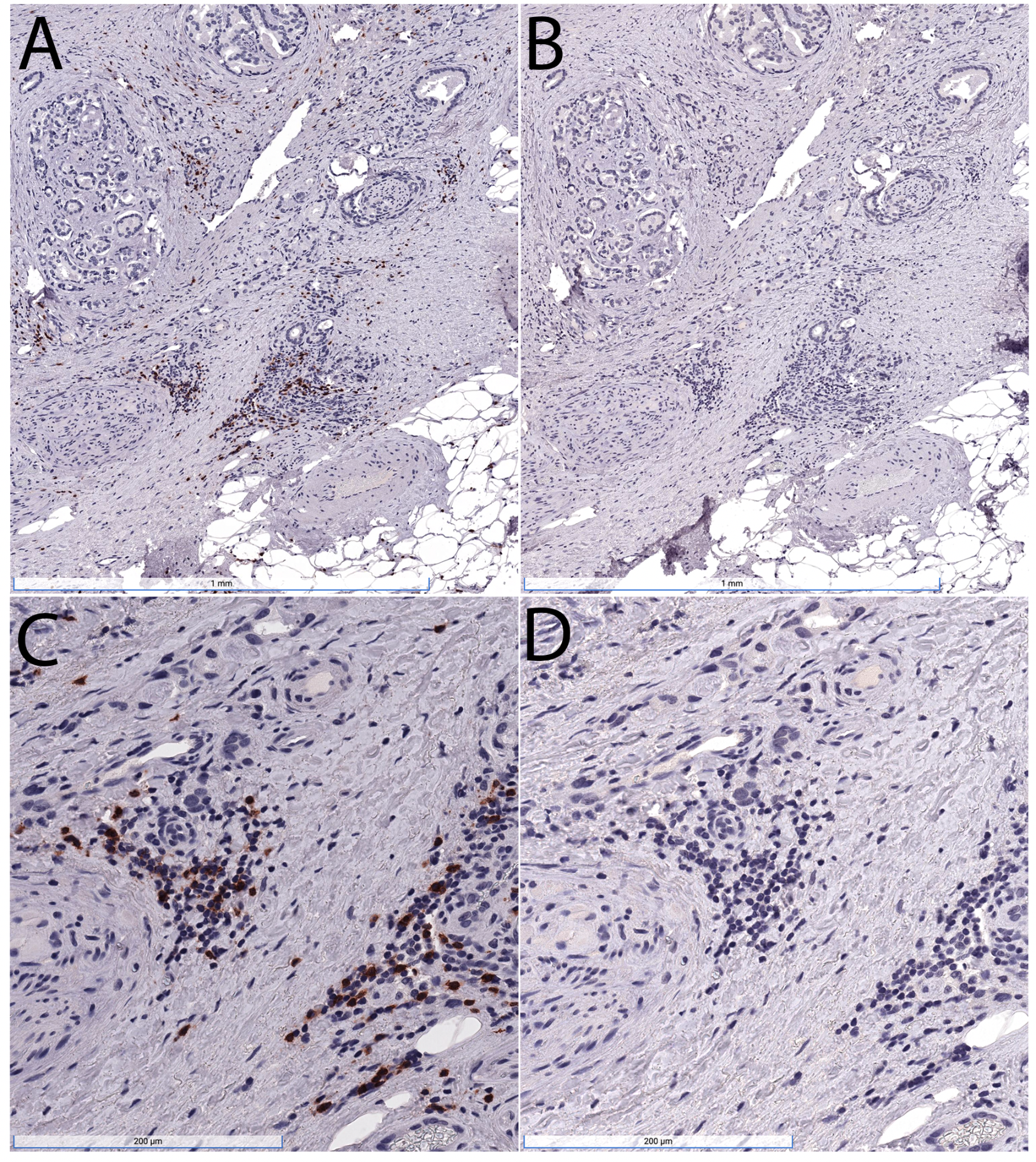

**Supplemental Figure 1.** **Removal of primary antibodies by microwave pretreatment between staining rounds**.  Tissue section from prostatectomy slide was stained for CD8 as in materials and methods. After staining and slide scanning (A,C), the cover slip was removed, AEC was dissolved in ETOH and the section was subjected to microwave antigen retrieval, followed by staining of the slide in an identical manner but with the secondary anti-mouse antibody only (B,D). Note the absence of the AEC signal with secondary only staining. Region of prostatic adenocarcinoma shown with CD8 cell infiltrate predominantly in the stroma.

**Supplemental Figure 2**

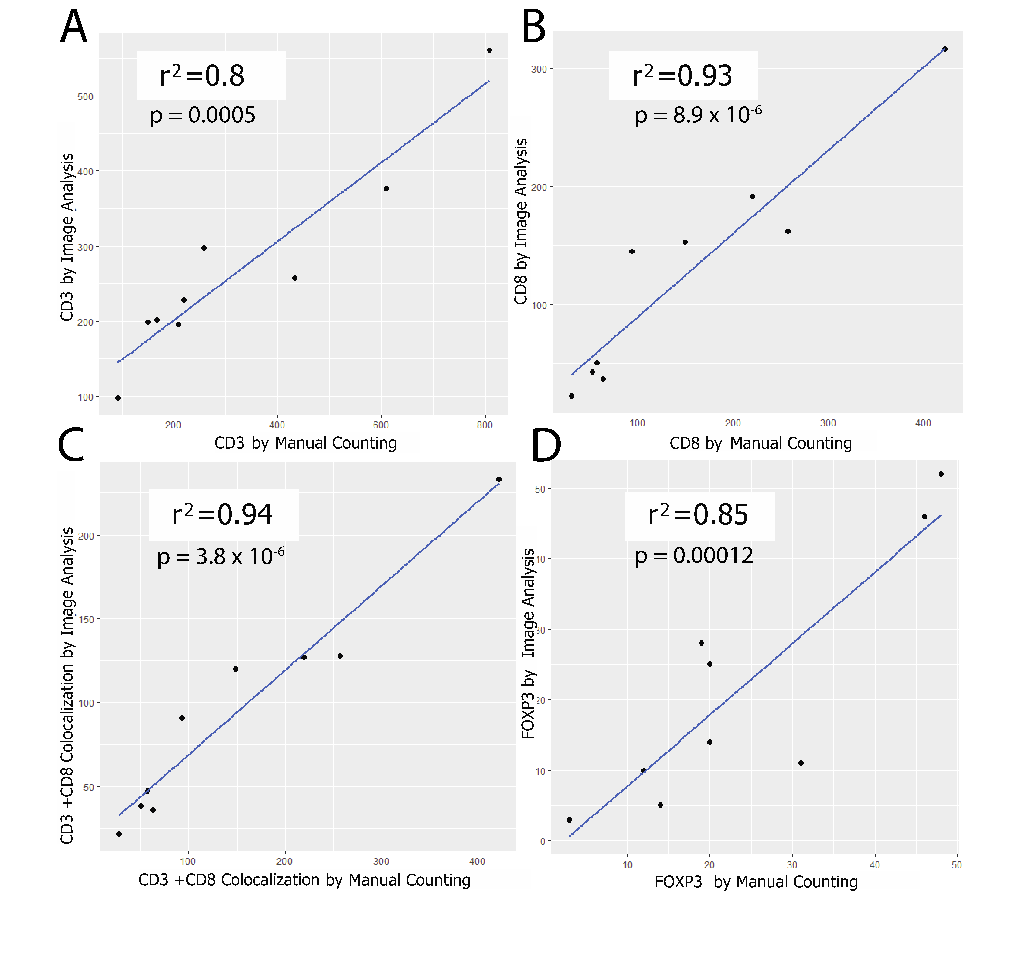

**Supplemental Figure 2.** **Validation of cell counting by image analysis**.  We compared cell counts using phenotypes defined in HALO by image analysis versus manual cell counting for several regions of interest across different standard slides from prostatectomies stained with the multiplex assay.  r^2^  and *P*-values are shown for the linear regression analysis.

**Supplemental Figure 3**

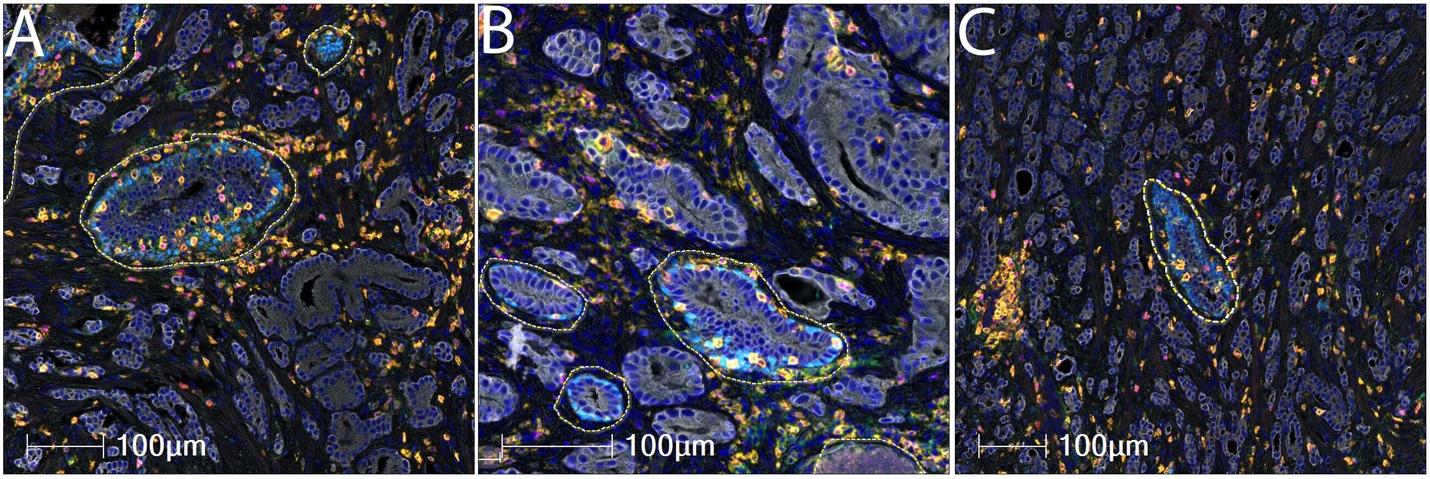

**Supplemental Figure 3.** **Annotations to exclude benign glands.** In many cases such as this one, adenocarcinoma glands infiltrate between pre-existing benign glands. In this case this is clearly seen in this medium power view in which the benign glands contain a layer of p63 positive basal cells. The benign glands are circled here for exclusion using a dashed yellow line. After benign glands are excluded the larger overall region of tumor is referred to as the benign-eliminated tumor region of interest (BET-ROI).

**Supplemental Figure 4**

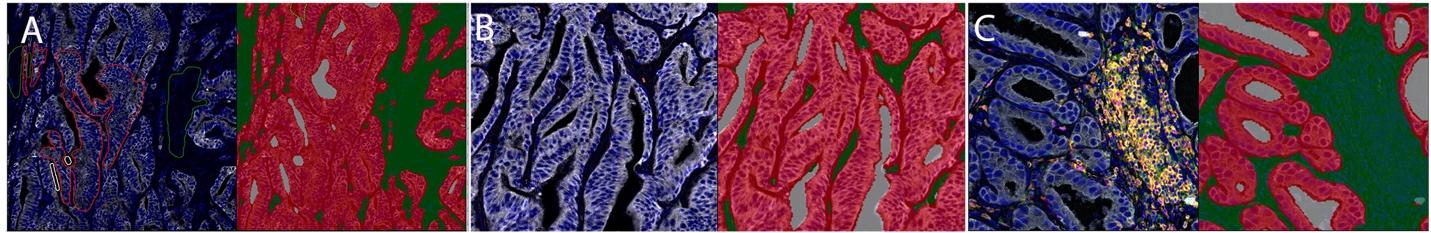

**Supplemental Figure 4**.  **Segmentation of epithelial and stromal compartments using HALO Classifier.**  **a** Left panel is pseudocolored IHC image and right shows results of the training in which the epithelium is shown in red,  the stroma in green and lumens in gray. **b** and **c** are two more examples of the results of using this classifier.

**Supplemental Table 1a. Antibodies, Pretreatment and Secondary Antibody Details**

| **Antigen** | **Source** | **Species** | **Clone** | **Dilution** | **Incubation Conditions** | **Antigen Retrieval** | **Secondary kit** | **Order in Panel** |
| --- | --- | --- | --- | --- | --- | --- | --- | --- |
| PD1 | Abcam | Mouse Monoclonal | NAT105 | 1:200 | ON* 4 °C | TR*** | UltraVision Quanto (Leica) | 1 |
| CD3 | DAKO | Rabbit | Polyclonal | 1:600 | 45 min RT** | Citrate | PowerVision+ (Leica PV6119) | 2 |
| CD8 | DAKO | Mouse Monoclonal | C8/144B | 1:200 | 45 min RT | Citrate | PowerVision+ (Leica PV6114) | 3 |
| CD4 | Abcam | Rabbit Monoclonal | EPR6855 | 1:2000 | 45 min RT | Citrate | PowerVision+ (Leica PV6119) | 4 |
| FOXP3 | eBioscience | Mouse Monoclonal | 236A/E7 | 1:250 | ON 4 °C | TR | PowerVision+ (Leica PV6114) | 5 |
| p63 | Cell Signaling Technology | Rabbit Monoclonal | D2K8X | 1:500 | 45 min RT | Citrate | PowerVision+ (Leica PV6119) | 6 |
| CK8 | Abcam | Rabbit Monoclonal | EP1628Y | 1:500 | 45 min RT | Citrate | PowerVision+ (Leica PV6119) | 7 |

*ON: overnight; **RT: Room temperature; ***TR (DAKO Target Retrieval Solution – S170084-2))

**Supplemental Table 1b. Cell Phenotypes**

| **Number** | **Cell types** | **Phenotype** |
| --- | --- | --- |
| 1 | T Cells All CD3+ | CD3+ |
| 2 | CD8+ T Cells All | CD3+CD8+ |
| 3 | CD4+ CD4+ Cells All | CD3+CD4+ |
| 4 | Regulatory T Cells | CD3+CD4+Foxp3+ |
| 5 | CD8+ T Cells - PD1 Positive | CD3+CD8+PD1+ |
| 6 | CD8+ T Cells - PD1 Negative | CD3+CD8+PD1- |
| 7 | CD4+ Cells - PD1 Positive | CD3+CD4+PD1+ |
| 8 | CD4+ Cells - PD1 Negative | CD3+CD4+PD1- |

**Supplemental Table 2. Pathology of Cases.**

| **Pt Num** | **Age** | **Race** | **Margins** | **Grade Group** | **Gleason Primary** | **Gleason Secondary** | **Gleason Sum** | **Gleason Tertiary** | **P Stage*** |
| --- | --- | --- | --- | --- | --- | --- | --- | --- | --- |
| 1 | 50-55 | W | Negative | 2 | 3 | 4 | 7 |  | T3BN0MX |
| 2 | 60-65 | O | Positive, NOS | 5 | 4 | 5 | 9 | 3 | T3BN0MX |
| 3 | 45-50 | W | Negative | 3 | 4 | 3 | 7 |  | T2N0MX |
| 4 | 40-45 | B | Positive, NOS | 4 | 3 | 5 | 8 | 4 | T3AN0MX |
| 5 | 50-55 | W | Negative | 5 | 4 | 5 | 9 |  | T2N0MX |
| 6 | 65-70 | W | Positive, NOS | 3 | 4 | 3 | 7 | 5 | T2XN0MX |
| 7 | 50-55 | W | Negative | 2 | 3 | 4 | 7 |  | T2N0MX |
| 8 | 60-65 | O | Negative | 5 | 4 | 5 | 9 |  | T3AN0MX |
| 9 | 60-65 | W | Negative | 3 | 4 | 3 | 7 |  | T3AN0MX |
| 10 | 60-65 | W | Negative | 2 | 3 | 4 | 7 |  | T2N0MX |
| 11 | 65-70 | W | Negative | 2 | 3 | 4 | 7 | 4 | T2NXMX |
| 12 | 55-60 | W | Positive, Moderate | 3 | 4 | 3 | 7 |  | T3AN0MX |
| 13 | 65-70 | W | Negative | 3 | 4 | 3 | 7 | 5 | T3AN0MX |
| 14 | 60-65 | W | Negative | 2 | 3 | 4 | 7 |  | T3AN0MX |
| 15 | 45-50 | W | Negative | 5 | 4 | 5 | 9 |  | T3BN1MX |

P Stage is the pathological stage at radical prostatectomy using the American Joint Committee on Cancer Staging 2007.

**Supplemental Table 3. T cell Densities by ERG in Total Tumor (PTEN Loss and PTEN Intact Combined)**

|  |  | **ERG Status** | |  |
| --- | --- | --- | --- | --- |
| **Cell Types** |  | **Negative** | **Positive** | ***p*** |
| T Cells All CD3+ | Median | 423.6 | 438.7 | 0.251 |
|  | Mean | 358.7 | 630.6 |  |
|  | SD | 156 | 422 |  |
| CD8+ T Cells All | Median | 125.8 | 121.4 | 0.475 |
|  | Mean | 115.2 | 252.3 |  |
|  | SD | 50.8 | 225.7 |  |
| CD4+ Cells All | Median | 194.8 | 343.3 | 0.0027 |
|  | Mean | 193.6 | 434.5 |  |
|  | SD | 80.5 | 213.4 |  |
| Regulatory T Cells | Median | 24.3 | 42 | 0.199 |
|  | Mean | 28.6 | 51.6 |  |
|  | SD | 22.3 | 38.7 |  |
| CD8+ T Cells – PD1 Positive | Median | 111 | 103.3 | 0.475 |
|  | Mean | 96.3 | 210.2 |  |
|  | SD | 40 | 189.4 |  |
| CD8+ T Cells – PD1 Negative | Median | 14.4 | 31.1 | 0.087 |
|  | Mean | 18.9 | 42 |  |
|  | SD | 14 | 37.1 |  |
| CD4+ Cells – PD1 Positive | Median | 162.6 | 265.4 | 0.0043 |
|  | Mean | 151.2 | 331.6 |  |
|  | SD | 151.2 | 201.4 |  |
| CD4+ Cells – PD1 Negative | Median | 34.9 | 102.3 | 0.007 |
|  | Mean | 42.4 | 103 |  |
|  | SD | 26.9 | 26.8 |  |

**Supplemental Table 4. T Cell Density by ERG Status**

| **Cell Type** | | ***p* PTEN Intact** | | | ***p* PTEN loss** | |
| --- | --- | --- | --- | --- | --- | --- |
| T Cells All CD3+ | | | 0.48 | | | 0.15 |
| CD8+ T Cells All | | | 0.78 | | | 0.32 |
| CD4+ Cells All | | | 0.39 | | | 0.01 |
| Regulatory T cells | | | 0.57 | | | 0.89 |
| CD8+ T Cells - PD1 Positive | | | 0.78 | | | 0.391 |
| CD8+ T Cells - PD1 Negative | | | 0.251 | | | 0.39 |
| CD4+ Cells - PD1 Positive | | | 0.48 | | | 0.032* |
| CD4+ Cells - PD1 Negative | | | 0.12 | | | 0.007* |

*P-*values reflect comparisons of ERG positive vs. ERG Negative, using Kruskal-Wallis equality-of-populations rank test. N=6 ERG positive and N=7 ERG negative cases.
